# Mental health of healthcare workers in England during the COVID-19 pandemic: a longitudinal cohort study

**DOI:** 10.1101/2022.06.16.22276479

**Authors:** Danielle Lamb, Rafael Gafoor, Hannah Scott, Ewan Carr, Sharon A.M Stevelink, Rosalind Raine, Matthew Hotopf, Neil Greenberg, Siobhan Hegarty, Ira Madan, Paul Moran, Richard Morriss, Dominic Murphy, Anne Marie Rafferty, Scott Weich, Sarah Dorrington, Simon Wessely

## Abstract

**Objective:** To examine variations in impact of the COVID-19 pandemic on the mental health of all types of healthcare workers (HCWs) in England over the first 17 months of the pandemic.

**Method:** We undertook a prospective cohort study of 22,501 HCWs from 18 English acute and mental health NHS Trusts, collecting online survey data on common mental disorders (CMDs), depression, anxiety, alcohol use, and PTSD, from April 2020 to August 2021. We analysed these data cross-sectionally by time period (corresponding to periods the NHS was under most pressure), and longitudinally. Data were weighted to better represent Trust population demographics.

**Results:** The proportion of those with probable CMDs was greater during periods when the NHS was under most pressure (measured by average monthly deaths). For example, 55% (95%CI 53%, 58%) of participants reported symptoms of CMDs in April-June 2020 versus 47% (95%CI 46%, 48%) July-October 2020. Contrary to expectation, there were no major differences between professional groups (i.e. clinical and non-clinical staff). Younger, female, lower paid staff, who felt poorly supported by colleagues/managers, and who experienced potentially morally injurious events were most at risk of negative mental health outcomes.

**Conclusion:** Among HCWs, the prevalence of probable CMDs increased during periods of escalating pressure on the NHS, suggesting staff support should be increased at such points in the future, and staff should be better prepared for such situations via training. All staff, regardless of role, experienced poorer mental health during these periods, suggesting that support should be provided for all staff groups.

**Key messages:** *What is already known on this topic:* Existing evidence about the mental health of healthcare workers (HCWs) through the COVID-19 pandemic comes mainly from cross-sectional studies using unrepresentative convenience samples, typically focussing on clinical staff rather than all HCWs. Such studies show high prevalence of symptoms of mental disorders, but the strength of this evidence is uncertain.

*What this study adds:* Using a defined sampling frame, with longitudinal, weighted data, we show that during periods of greater pressure on the NHS (as indicated by average monthly national COVID-19 death rates), prevalence of mental disorder symptoms increased, and, importantly, that this effect was seen in non-clinical as well as clinical staff.

*How this study might affect research, practice or policy:* These findings indicate that provision of support for HCWs should not only focus on those providing clinical care, but also on non-clinical staff such as porters, cleaners, and administrative staff, and additional support should be provided during higher pressure periods. Better preparation of staff for such situations is also suggested.

## Introduction

Much has been written about the pressure that the COVID-19 pandemic has placed on the NHS. Health Care Workers (HCWs) are defined here as anyone working in a healthcare setting, including clinical and non-clinical roles. Those in non-clinical roles (e.g. porters, cleaners, receptionists) and non-patient-facing roles (e.g. HR, finance departments, administration) are as integral to the functioning of the NHS as clinical staff, and yet have largely been excluded from existing research. Evidence from the NHS Staff Survey shows that the proportion of NHS staff feeling unwell due to work-related stress rose from 40% in 2019 to 44% in 2021, though it is worth noting that there has been a steady increase since 2016, when 36% of staff reported this (NHS England, 2021).

UK data indicates that overall population mental health deteriorated during the pandemic, with statistically significantly higher prevalence of psychological distress in April 2020 than in previous years (Daly et al., 2020; Pierce, Hope, et al., 2020). However, there is conflicting evidence about whether this undoubted increase in population prevalence is even more marked in HCWs, as might be expected given the additional pressures placed on them by the pandemic (Pierce, Hope, et al., 2020). Systematic reviews report pooled prevalence estimates for symptoms of anxiety of 30% (95%CI 24, 37), of depression of 31% (95 %CI 26, 37), and of Post-Traumatic Stress Disorder (PTSD) of 20% (95%CI 10, 33) (Marvaldi et al., 2021). These estimates are in line with levels of mental health symptoms found in HCWs in previous epidemics and pandemics (Preti et al., 2020).

Existing evidence suggests that being female and younger in age are risk factors for experiencing worse mental health during the pandemic (Daly & Robinson, 2021). There is also emerging evidence about associations between poor mental health and experiencing events that conflict with one’s own values (Williamson et al., 2021). These are known as ‘potentially morally injurious events’ (PMIEs), and in healthcare settings can include accidental or deliberate negligence, witnessing unethical behaviour and failing to intervene, or betrayal by trusted others (e.g. managers not providing appropriate support or resources), events which the context of the pandemic in addition to existing pressures may have made more likely. Our own and other research has also indicated the importance of support from peers and managers, access to personal protective equipment (PPE), and associations between job role and setting with mental health outcomes (Greenberg et al., 2021; Lamb, Gnanapragasam, et al., 2021; Wanigasooriya et al., 2021).

To date, much research has considered only clinical staff, limiting generalisability, and nearly all previous studies (finding high levels of poor mental health among HCWs) have relied on cross-sectional data (Greenberg et al., 2021). Many of these studies also rely on convenience samples, with no clear sampling frame and the attendant risk of selection bias (Gnanapragasam et al., 2021; Lamb et al., 2020; Pierce, McManus, et al., 2020). The NHS CHECK study addresses these limitations by including all HCW staff in participating Trusts (clinical and non-clinical), and providing longitudinal data.

### Aims and hypotheses

The primary aim of this paper is to examine changes in mental health outcomes in HCWs during the first 17 months of the COVID-19 pandemic, from April 2020 to August 2021 inclusive. Our main research question was: What are the changes in mental health outcomes over time adjusting for potential covariates? We hypothesised non-linear time trends that track pressure on the NHS, with higher levels of reported symptoms in the earlier months (April to June 2020), with prevalence reducing through the summer (July to October 2020), before rising again going into the autumn and winter (November 2020 to March 2021), before falling again through spring and summer (April to August 2021).

The secondary aims of this paper were to investigate differences associated with mental health outcomes in NHS staff including: i) socio-demographic characteristics; ii) work setting; iii) job role; iv) occupational factors (e.g. access to PPE); v) organisational support available; and vi) exposure to PMIEs. Our secondary research question was: What factors are associated with poorer mental health? We hypothesised higher prevalence of negative mental health outcomes among women, younger workers, and those in racial and ethnic minority groups, as well as those with inadequate access to PPE, those lacking support from colleagues/managers, and those working in Intensive Care Units (ICU) and Accident & Emergency (A&E) settings (Lamb, Gnanapragasam, et al., 2021). We also hypothesised that those who report greater exposure to PMIEs will report poorer mental health (Williamson et al., 2018).

## Methods

### Study design and participants

The target population for NHS CHECK was all staff (clinical and non-clinical working in 18 participating NHS Trusts. The baseline data collection period was April 2020 to January 2021. Follow up data were collected approximately six months after each individual completed the baseline survey, beginning in November 2020 and ending in August 2021. Collecting data at just two time points for each participant balanced our need for longitudinal data against the burden on staff (Gnanapragasam et al., 2021).

### Procedures

Trusts were invited to participate via direct emails to senior leadership teams. Trusts were selected to offer diversity in geographical location, urban and rural settings, acute and mental health Trusts (for more details see Lamb, Greenberg, et al., 2021). Participating Trusts circulated emails explaining and promoting the study to all eligible staff via existing group email lists. A dedicated NHS CHECK recruitment email was sent by senior Trust management with a link to the study website (www.nhscheck.org), with variations of this email sent repeatedly during baseline recruitment. We also used existing staff support teams/leads, chief nursing officers, medical directors, occupational health departments, trade union representatives, and wellbeing hub users to promote the study. NHS CHECK was brought up during team briefings, included in Trust newsletters, news items on Trust intranet websites, closed social media groups and advertised via screen savers on Trust computers. We fed back recruitment data to Trusts on a weekly basis, and provided incentives such as prizes (e.g. coffee machines) for Trusts with the highest proportions of recruitment. All participants who gave consent to be contacted again were entered into a prize draw to win one of 10 £50 and 10 £250 gift vouchers.

### Data collection and materials

The NHS CHECK Patient and Public Involvement (PPI) advisory group co-developed a questionnaire, with the acceptability of the questions, materials, and procedures checked with a small group of psychologists, managers, administrators, security staff, intensivists, and trainees. To participate, after reading an information sheet, staff completed an online consent form, and the online questionnaire then took around 5-10 minutes, with an option to complete a longer additional questionnaire (10-15 minutes). The NHS CHECK baseline questionnaire was launched on the 24^th^ of April 2020, 5 weeks after the initial lockdown in the UK began (23/03/20). Baseline recruitment closed on 15/01/21, with the long roll-out period due to additional sites joining the study as it progressed (see Lamb, Greenberg, et al., 2021 for details). The six-month follow up survey was sent via email to participants six months after their baseline survey completion date, with a four-week window in which to complete the survey, and data collection closing on 15/08/2021.

#### Surveys

Surveys collected information on participants’ socio-demographic characteristics, their occupational role and context, COVID-19 experiences, and a range of validated measures (Lamb, Greenberg, et al., 2021).

The primary outcome was prevalence of probable common mental disorders (CMDs), ascertained using the 12-item General Health Questionnaire (GHQ-12). The GHQ scoring method was used (where each item is scored 0-0-1-1, resulting in a total score of 0-12 for the scale), with a cut-off score of 4 or more indicating ‘caseness’ of a CMD (indicating increased probability of experiencing a recognised mental disorder) (Goldberg & Hillier, 1979).

Secondary outcomes were anxiety, assessed using the Generalised Anxiety Disorder scale (GAD-7) with a cut-off of ≥10 indicating caseness (Spitzer et al., 2006), and depression, using the Patient Health Questionnaire (PHQ-9) with a cut-off of ≥10 indicating caseness (Kroenke et al., 2001). The Alcohol Use Disorder Identification Test (AUDIT-C) assessed problem drinking, with a cut-off of ≥8 indicating caseness (Babor et al., 2001). The Post-Traumatic Stress Disorder checklist (PCL–6) assessed PTSD, with a cut-off of ≥14 indicating probable caseness (Lang & Stein, 2005). The Moral Injury Events Scale (MIES) was used to collect data on exposure to potentially morally injurious events (PMIEs) (Nash et al., 2013), with endorsement of one or more item indicating exposure (i.e. selecting moderately/strongly agree). At each time point participants were invited to complete a ‘short’ survey, with the option to continue on and complete a ‘long’ survey in addition. See supplementary table S1 for information about which measures were used in which survey.

### Statistical analysis

#### Data cleaning and weighting

Where participants erroneously completed the survey more than once at baseline, or at six-months, duplicate responses (identified by identical email address) were dropped, using the following process: static data (i.e. demographics such as age, sex, ethnicity, role) were imputed across duplicate responses; the most complete response containing the primary outcome measure (GHQ-12) was retained; where duplicate responses were equally complete, the earliest, most complete response was retained.

Participating Trust HR departments provided population-level demographic data as of April 2020, including the number of employees and the age, sex, and ethnicity composition of the workforce, as well as a breakdown by job role. From this, we calculated a response rate for each Trust. Response weights were generated using a raking algorithm based on age, sex, ethnicity, and role, using the full baseline cohort. For weighting purposes only, missing demographic data were imputed using the 5^th^ nearest neighbour (kNN) algorithm. Missingness was no more than 5% for age, sex, ethnicity, and role at baseline data collection. Individual person level strata were weighted by Trust size as well as composition of the four main proposed prognostic factors of ethnicity, categorical age, role and sex. Post strata and post weight specifications were included but no finite population correction was made.

#### Partitioning of data by time-period

Given the extended data collection periods at baseline and six-months, we identified ‘burden periods’ – periods of time when pressures and burden on NHS Trusts and staff were higher and lower. For example, in line with previous research (Patel et al., 2021), we identified burden as: high early in the pandemic (T1: April-June 2020), when little was known about COVID-19 and swift changes were needed in healthcare settings and in general life; while burden was lower by the summer/early autumn of 2020 (T2: July-October 2020), when the initial wave of infections lessened and lockdown restrictions eased; before increasing again over the winter months (T3: November 2020-March 2021); and then reducing again during spring/summer 2021 (T4: April-August 2021). These time periods correspond to periods with higher and lower COVID-19 death rates: T1 COVID-19 deaths = 53,389; T2 COVID-19 deaths = 7,157; T3 COVID-19 deaths = 88,211; T4 COVID-19 deaths = 6,969 (Office for National Statistics, 2022). This enabled us to explore outcomes and associated factors in more detail and compare our results to other research conducted during similar time periods.

#### Descriptive analyses

We explored the data in several stages. First, we described the characteristics of those completing the baseline survey(s), and those completing the six-month survey(s). We then assessed the representativeness of the sample to the HR population data, using frequencies and percentages for categorical variables, mean and standard deviation for continuous variables. Next, we described the demographic differences between participants completing both the short and long baseline surveys versus those completing only the short survey, at baseline and six-month surveys. Finally, we summarized the weighted prevalence and mean scores of the primary (GHQ-12) and secondary outcomes (GAD-7, PHQ-9, AUDIT-C, PCL-6) cross-sectionally by burden periods (T1-T4).

#### Cross-sectional analyses

As data were clustered by Trust, we carried out multilevel multivariable logistic regressions to explore factors associated with the primary and secondary outcomes, by burden periods. The factors included were decided on *a priori*, based on existing evidence and team discussions: age category; sex; ethnicity; relationship status; job role; pay grade; job setting; had good access to PPE; felt supported by colleagues; felt supported by managers; redeployed; experienced moral injury. We modelled outcomes adjusting for all other variables in the model.

#### Longitudinal analyses

Changes over time were modelled for the outcome variables (GHQ-12, GAD-7, PHQ-9, and PCL-6). Analyses were controlled for the following pre-determined confounders – ethnicity (using the ONS five top-level categories), sex, categorical age, and role (clinical/non-clinical) at baseline. Time was modelled using fractional polynomials within hierarchical generalized linear models to account for clustering of participants at Trust level. Model diagnostics were assessed visually using post estimation plots of residuals and caterpillar plots. To enable cross model comparison, the covariates from the best fitting model with GHQ-12 as an outcome (using Bayesian Information Criteria) were applied to the analyses of the other outcomes. Complete case analyses were performed and responses with missing data after kNN(5) imputation were dropped. Akaike and Bayesian Information Criteria (AIC/BIC) were used in deciding whether to introduce an interaction term. Where the interaction term is deemed significant, stratified results are presented. An absolute difference of 2 points in both AIC and BIC were taken as evidence against the null hypothesis that an interaction term did not improve the model fit.

All analyses were conducted using Stata v 16.1, R v4.1.1 & R Studio v1.4.1717.

### Ethical approval

Ethical approval for the study was granted by the Health Research Authority (reference: 20/HRA/210, IRAS: 282686) and local Trust Research and Development approval. The study was approved as having Urgent Public Health Status in August 2020.

## Results

The total sample size, before data cleaning, was 26,486. Once completely empty responses and duplicate responses were dropped the total sample size was 23,462. As specified in the protocol (Lamb, Greenberg, et al., 2021), any Trust with <5% response rate was dropped. After dropping the one Trust with a response rate <5% at baseline, and respondents who did not report a Trust, the final sample size was 22,501 (total summed Trust population=139,037), which represents an overall response rate of 16% (range: 5% to 55%). At six-months, the total sample size was 10,671, which reflects an overall response rate of 8% (Trust range: 3%-20%).

### Descriptive analyses

#### Cohort demographics

The demographic composition of the sample can be seen in **Error! Not a valid bookmark self-reference**., which shows the total number of respondents in each category, with total unweighted and weighted proportions given, at baseline and six-months. At baseline, just under half completed only the short survey (n=9,987; long survey n=12,514). Similarly, at six-months around half completed only the short survey (n=5,333; long survey n=5,338). At baseline, women were statistically significantly more likely to complete both surveys than men, as were those in older age categories, White participants, those in a relationship, those who had not been redeployed, those who felt supported by colleagues and managers, and those who were nurses or in other clinical or non-clinical roles (compared to doctors). At six-months, women were statistically significantly more likely to complete both surveys, as were those in older age categories, those in community or non-patient-facing settings, those with adequate access to PPE, and those on lower pay grades (see supplementary table S2).

#### Prevalence of outcomes

As can be seen in Table 2, in line with our hypotheses, the proportions of participants meeting cut-off score for common mental disorders tended to be higher at T1 and T3 (when burden on the NHS was higher) than at T2 and T4. For example, at the start of the pandemic T1, 55% (95%CI 52, 58) of participants met cut-off on the GHQ-12, with the proportion being lower at T2 as pressure and restrictions eased (47%, 95%CI 46, 48), before rising again as pressure increased through the winter T3 (55%, 95%CI 53, 56), and reducing again through spring and summer T4 (51%, 95%CI 48, 54). This pattern was similar for PTSD and depression (though the latter remained high at T4), while the proportion meeting cut-off for anxiety rose slightly from T1 (21%, 95%CI 19, 23) to T2 (22%, 95%CI 21, 24), and then rose again at T3 (25%, 95%CI 24, 27) and T4 (25%, 95%CI 22, 28). The proportions meeting cut-off for alcohol misuse remained relatively stable over time. The proportion of participants meeting cut-off score on each measure, together with the total number of COVID-19 deaths, are shown by time period in Figure 1.

**Table 1.** Descriptive characteristics of the cohort at baseline and six-month time points, with weighted and unweighted proportions given

| Characteristic | n | Baseline |  | Six-months |  |  |
| --- | --- | --- | --- | --- | --- | --- |
|  |  | Unweighted % | Weighted % | N | Unweighted % | Weighted % |
| Age (years) |  |  |  |  |  |  |
| ≤30 | 4,277 | 19 | 21 | 1,623 | 15 | 16 |
| 31-40 | 4,939 | 22 | 25 | 2,160 | 20 | 24 |
| 41-50 | 5,611 | 25 | 23 | 2,802 | 26 | 24 |
| 51-60 | 5,214 | 23 | 20 | 2,852 | 27 | 23 |
| ≥61 | 1,304 | 6 | 6 | 739 | 7 | 8 |
| Missing data | 1,156 | 5 | 5 | 495 | 5 | 5 |

| Characteristic | Baseline |  |  | Six-months |  |  |
| --- | --- | --- | --- | --- | --- | --- |
|  | n | Unweighted % | Weighted % | N | Unweighted % | Weighted % |
| Sex |  |  |  |  |  |  |
| Female | 18,067 | 80 | 74 | 8,656 | 81 | 75 |
| Male | 4,172 | 19 | 25 | 1,954 | 18 | 25 |
| Missing data | 262 | 1 | 1 | 61 | 1 | <1 |
| Relationship status |  |  |  |  |  |  |
| In a relationship | 16,449 | 73 | 72 | 7,953 | 75 | 25 |
| Single | 5,817 | 26 | 27 | 2,663 | 25 | 74 |
| Missing data | 235 | 1 | 1 | 55 | <1 | 1 |
| Ethnicity |  |  |  |  |  |  |
| White | 19,085 | 85 | 75 | 9,411 | 88 | 80 |
| Black/African/Caribbean /Black British | 989 | 4 | 9 | 335 | 3 | 6 |
| Asian/Asian British | 1,481 | 7 | 12 | 571 | 5 | 10 |
| Mixed/Multiple racial and ethnic groups | 546 | 2 | 1 | 238 | 2 | 1 |
| Other racial and ethnic minority groups | 207 | 1 | 2 | 81 | 1 | 2 |
| Missing data | 193 | 1 | 1 | 35 | 1 | 1 |
| Main role |  |  |  |  |  |  |
| Doctor | 1,653 | 7 | 10 | 757 | 7 | 10 |
| Nurse | 5,741 | 26 | 29 | 2,613 | 24 | 28 |
| Other clinical | 6,778 | 30 | 31 | 3,078 | 29 | 30 |
| Non-clinical | 7,977 | 35 | 29 | 4,117 | 39 | 31 |
| Missing data | 352 | 2 | 1 | 106 | 1 | 1 |
| Job setting |  |  |  |  |  |  |
| A&E | 336 | 1 | 3 | 153 | 1 | 2 |
| ICU/Anaesthetics | 799 | 4 | 5 | 423 | 4 | 5 |
| Other hospital | 12,899 | 57 | 66 | 6,288 | 59 | 66 |
| Community | 6,653 | 30 | 19 | 3,023 | 28 | 20 |
| Non-patient-facing | 1,049 | 5 | 4 | 585 | 6 | 5 |
| Missing data | 765 | 3 | 3 | 199 | 2 | 2 |
| Redeployed |  |  |  |  |  |  |
| Yes | 2,783 | 12 | 12 | 1,241 | 88 | 11 |
| No | 19,365 | 86 | 86 | 9,373 | 12 | 88 |
| Missing data | 353 | 2 | 2 | 57 | <1 | 1 |
| PPE access often/always |  |  |  |  |  |  |
| Yes | 17,049 | 76 | 77 | 8,081 | 76 | 78 |
| No | 1,708 | 7 | 9 | 793 | 7 | 8 |
| Missing data | 3,744 | 17 | 14 | 1,797 | 17 | 14 |
| Felt supported by colleagues |  |  |  |  |  |  |
| Yes | 19,970 | 89 | 88 | 9,728 | 91 | 91 |
| No | 1,683 | 7 | 8 | 794 | 8 | 8 |
| Missing data | 848 | 4 | 4 | 149 | 1 | 1 |
| Felt supported by manager |  |  |  |  |  |  |
| Yes | 18,143 | 81 | 79 | 8,856 | 83 | 81 |
| No | 3,484 | 15 | 17 | 1,654 | 15 | 17 |
| Missing data | 874 | 4 | 4 | 161 | 2 | 2 |
| Pay grade<sup>a</sup> |  |  |  |  |  |  |
| AfC 5 or below | 7,112 | 31 | 28 | 3,457 | 32 | 28 |

|  |  | Baseline |  |  | Six-months |  |  |
| --- | --- | --- | --- | --- | --- | --- | --- |
| Characteristic |  | n | Unweighted % | Weighted % | N | Unweighted % | Weighted % |
|  | AfC 6 or above (inc. medical pay scales) | 12,081 | 54 | 55 | 5,924 | 56 | 57 |
|  | Missing data | 3,308 | 15 | 17 | 1,290 | 12 | 15 |

a Pay scale is dichotomised at approximately the median national average wage in the UK, £30,472 (Smith, 2020), using the Agenda for Change (AfC) pay scales (NHS, 2020) and medical pay scales (Scavone, 2021).
Abbreviations: ICU – Intensive Care Unit; PPE – Personal Protective Equipment

**Table 2.**
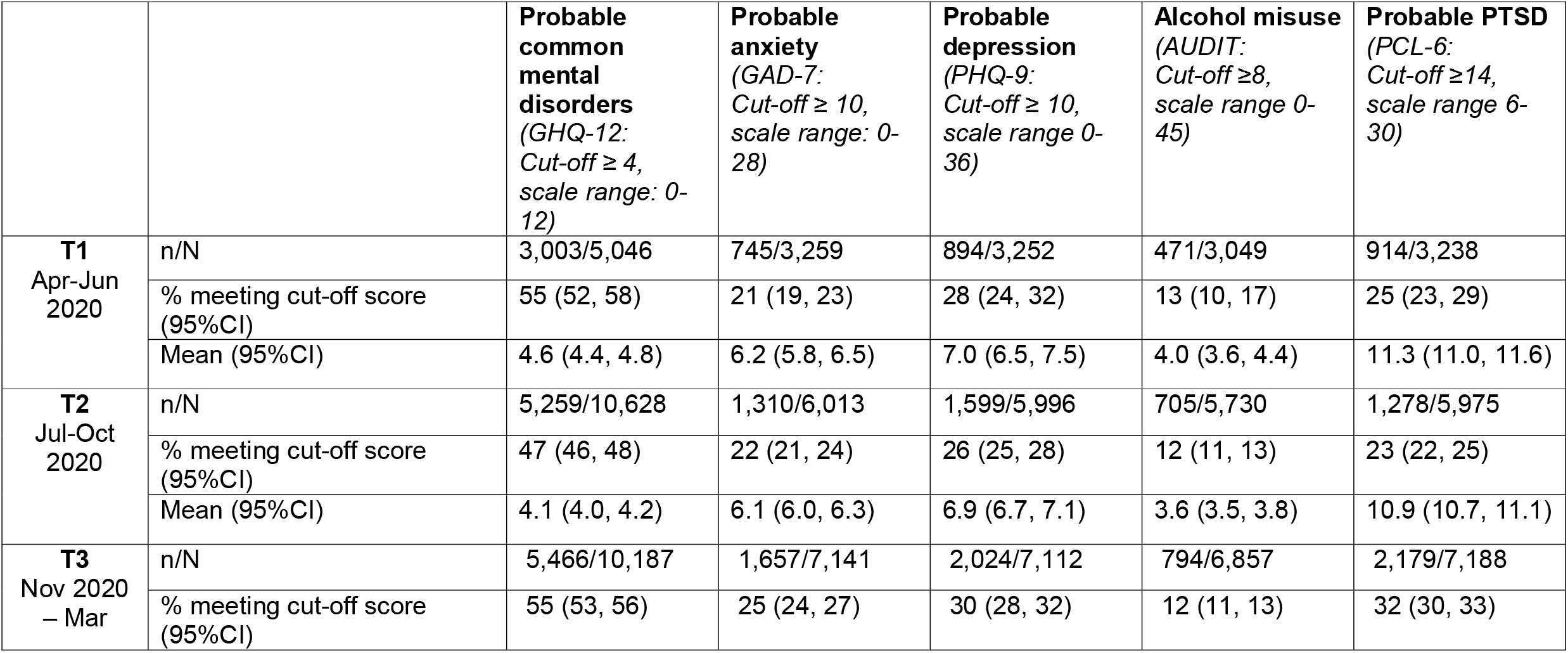
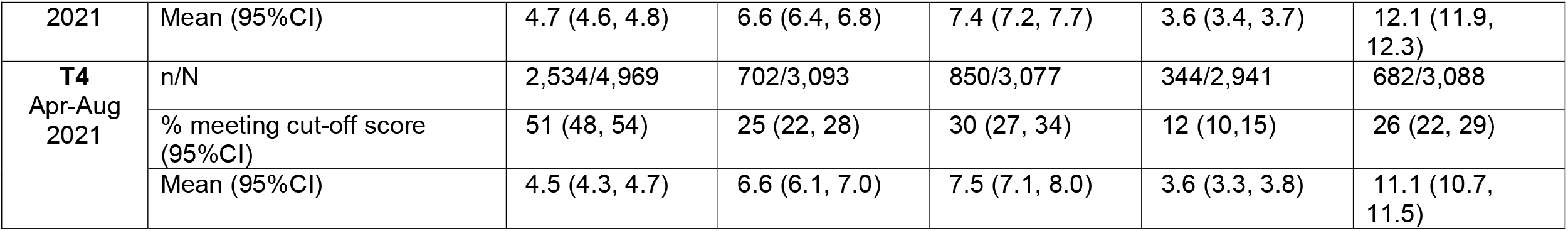
Prevalence of adverse mental health outcomes, by burden period

**Figure 1.**
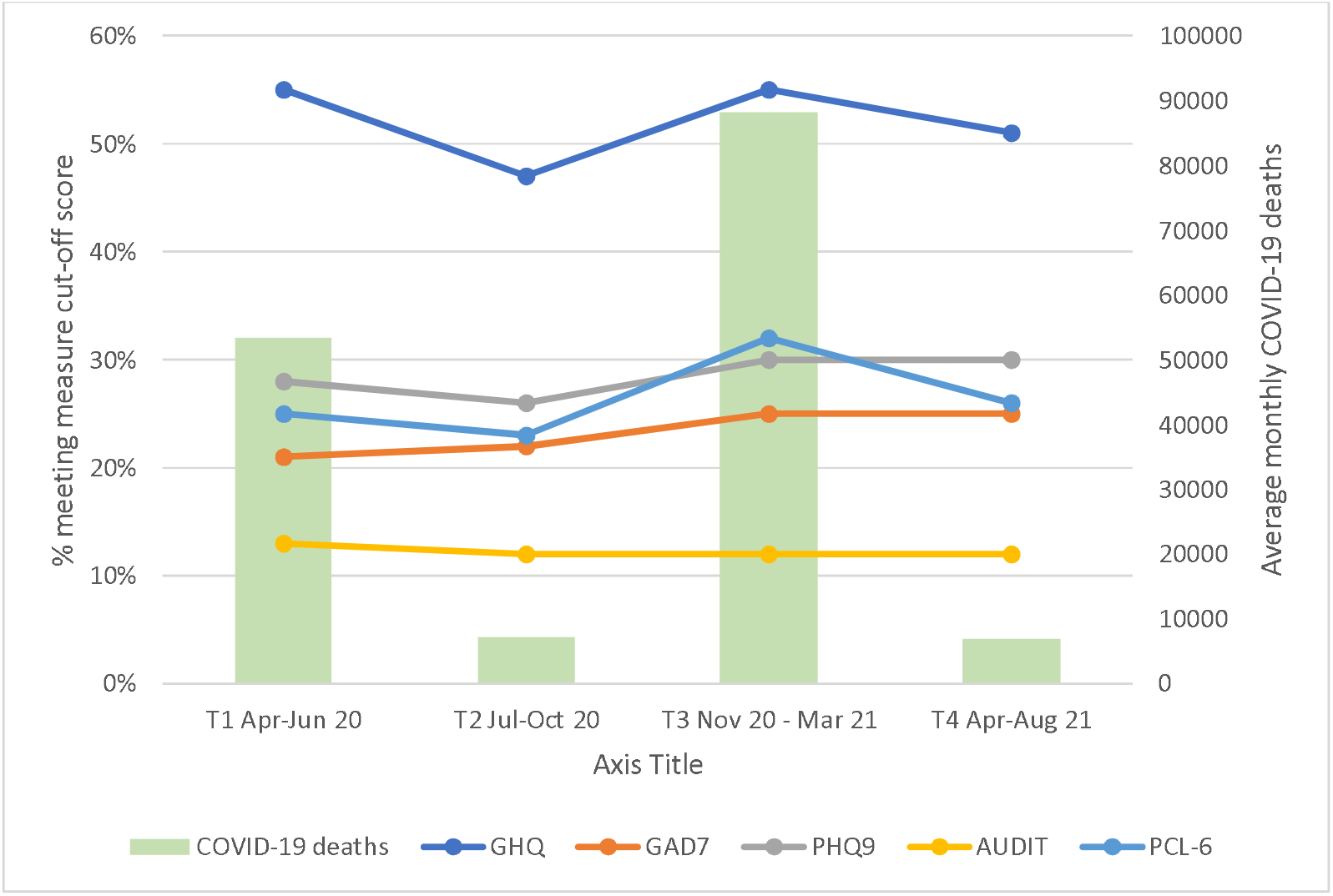
Proportions meeting cut-off score on each measure, with COVID-19 deaths, by time period

### Cross-sectional analyses

#### Common mental disorders (primary outcome)

The primary outcome was CMDs, as measured by the GHQ-12. As can be seen in Table 3, the factors associated with meeting cut-off on the GHQ-12 varied across time periods. The factors that appear to remain broadly consistent throughout the pandemic are that older participants were less likely than younger participants to meet cut-off, those who felt well supported by colleagues and managers were less likely to meet cut-off than those who did not feel supported, and those reporting higher exposure to morally injurious events were more likely to meet cut-off than those not experiencing moral injury. At points where the NHS was under more pressure (i.e. T1 and T3), women were more likely than men to meet cut-off, those who redeployed were less likely to meet cut-off than those not redeployed (T3), and Asian participants were less likely to meet cut-off than White participants. Those reporting mixed ethnicity were more likely than White participants to meet cut-off at T2, and those reporting other ethnicities were less likely than White participants to meet cut-off at T4. In terms of role, all staff groups were less likely to meet cut-off than doctors at T2, and nurses were more likely to meet cut-off than doctors at T3. In terms of job setting, those in all other areas were more likely to meet cut-off at T1 than A&E staff, while those in non-patient facing areas were less likely than A&E staff to meet cut-off at T3. Those with good access to PPE were less likely to meet cut-off at T1 than those without access.

**Table 3.**
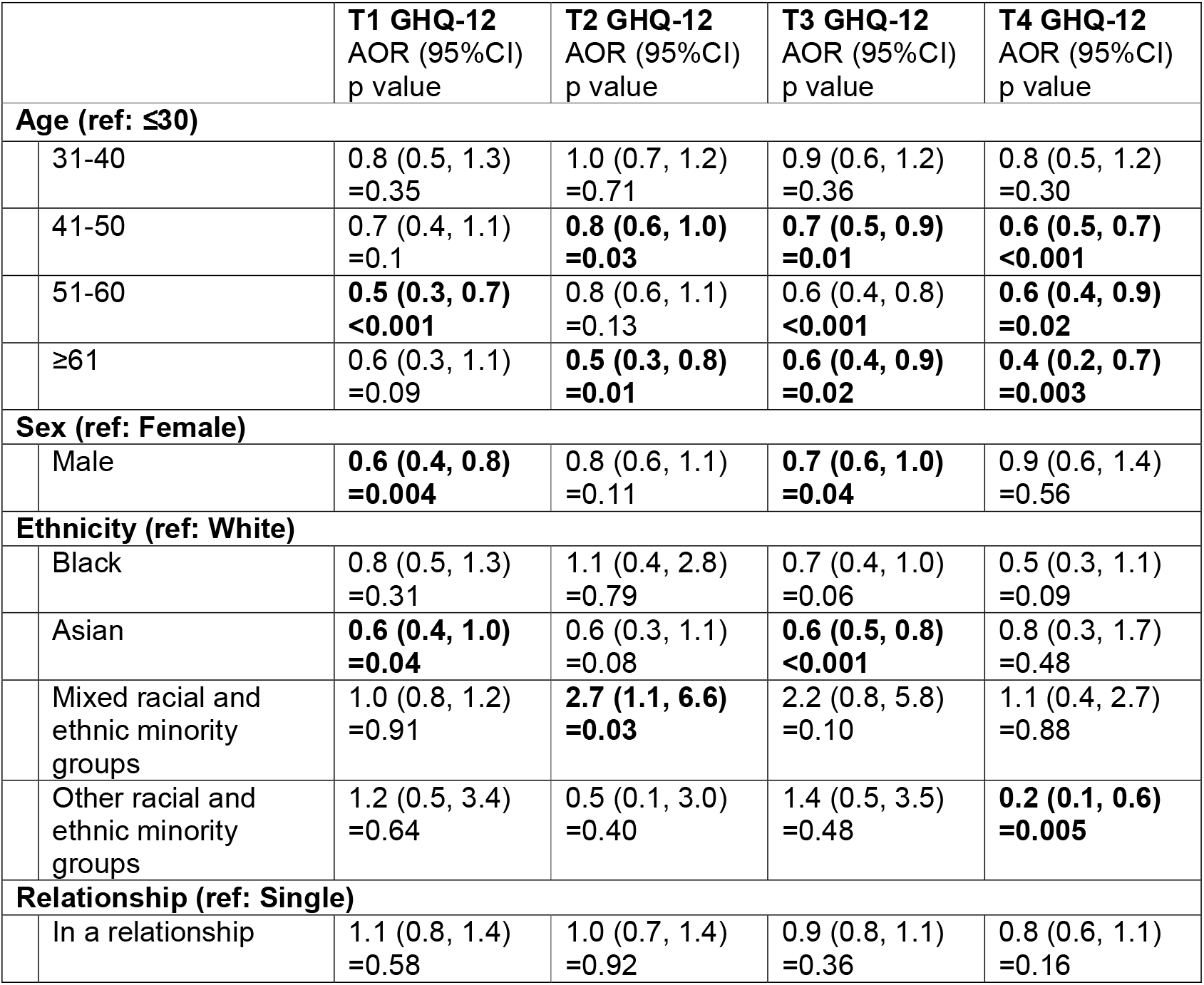
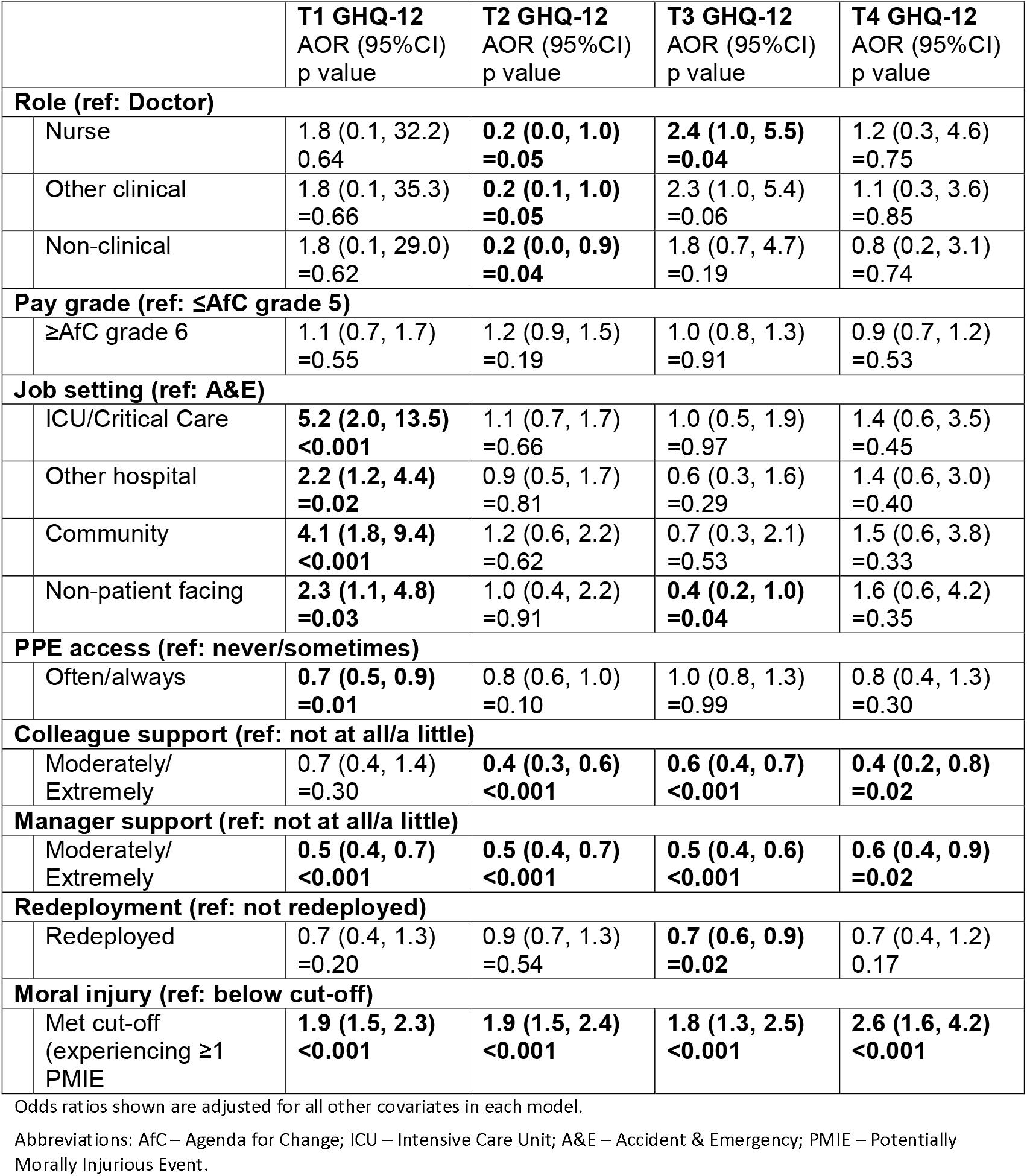
Weighted, multilevel, multivariable binary logistic regression models of factors associated with meeting GHQ-12 cut-off, by burden period

#### Secondary outcomes

The results of similar analyses for the secondary outcomes (anxiety, depression, alcohol misuse, and PTSD) are given in Supplementary tables S3, S4, S5, and S6, respectively.

As expected, across all time periods, those in older age categories were less likely to meet GAD-7 cut-off for probable anxiety disorders than those ≤30 (e.g. T3 51-60, AOR 0.4, 95%CI 0.3, 0.6, p<0.001). Also, as in the general population, in all periods except T1, men were less likely to meet cut-off than women (e.g.T4 AOR 0.9, 95%CI 0.5, 0.9, p=0.03), as were those in higher pay grades compared to those in lower pay grades (e.g. T3 AOR 0.7, 95%CI 0.5, 0.8, p<0.001). Across all time points except T4, those who felt well supported by their manager were less likely to meet cut-off than those who did not feel supported (e.g. T1 AOR 0.6, 95%CI 0.5, 0.8, p<0.001). A more novel and specific finding is that those who were exposed to higher levels of moral injurious events were more likely to meet cut-off than those who were not, across all time periods (e.g. T1 AOR 2.7, 95%CI 2.1, 3.5, p<0.001).

Much the same is true for probable depression. Across all time periods those in older age categories were less likely to meet PHQ-9 cut-off than those ≤30 years old (e.g. T1 41-50 AOR 0.7, 95%CI 0.4, 0.9, p=0.03). Across all time points other than T1, those in a relationship were less likely to meet cut-off than those not in a relationship (e.g. T4 AOR 0.7, 95%CI 0.5, 0.8, p<0.001). Those in higher pay grades were less likely to meet cut-off than those in lower pay grades, across all time points (e.g. T3 AOR 0.5, 95%CI 0.4, 0.7, p<0.001). Those with supportive colleagues and/or managers were less likely to meet cut-off, across all time points (e.g. T2 AOR 0.5, 95%CI 0.4, 0.8, p=0.01). Those who experienced moral injury were more likely to meet cut-off than those who did not, across all time periods (e.g. T2 AOR 2.4, 95%CI 2.0, 2.9, p<0.001).

Turning to alcohol use, men were consistently more likely than women to meet AUDIT cut-off, across all time points (e.g. T1 AOR 2.0, 95%CI 1.4, 2.9, p<0.001). Those of Black, Asian, or mixed racial or ethnic groups were less likely to meet cut-off than White participants, across all time points (e.g T2 AOR 0.3, 95%CI 0.1, 0.7, p=0.01).

Men were consistently less likely than women to meet PCL-6 cut-off for probable PTSD than women, across all time points (e.g. T3 AOR 0.7, 95%CI 0.5, 0.9, p=0.02). Apart from at T2, those on higher pay grades were less likely to meet cut-off than those on lower pay grades (e.g. T1 AOR 0.8, 95%CI 0.7, 0.9, p<0.001). Those working in ICU were more likely to meet cut-off than those in other job settings, at all time points except T4 (e.g. T3 2.9, 95%CI 1.5, 5.6, p<0.001). Those who were exposed to more morally injurious events were more likely to meet cut-off than those who did not, across all time periods (e.g. T4 AOR 3.5, 95%CI 2.6, 4.9, p<0.001).

### Longitudinal analyses

All results demonstrated evidence of improved model fit with an interaction term between the variables which coded for age categories and sex. Therefore, stratified analyses by sex are presented. Results model the odds ratios (ORs) of meeting cut-off on the GHQ-12, GAD-7, PHQ-9, AUDIT, and PCL-6 over the study period, for the predictor variables of age categories, ethnicity, and role, accounting for time. The analyses are hierarchical and therefore account for inter-person clustering (for repeated measures) as well as clustering by trust.

#### Common mental disorders (primary outcome)

There was no statistically significant evidence of a difference in the ORs of meeting GHQ-12 cut-off over the study period for those in Other or Black ethnic groups for either sex, however there was a significant reduction in the OR for those who from Asian versus White ethnic groups. Non-clinical staff had a reduced OR of meeting GHQ-12 cut-off versus clinical staff for both sexes. There were reductions across almost all age categories for both sexes in the OR of meeting GHQ-12 cut-off (but not significantly different for males aged 41yo to 50yo or for females ages 31yo to 40yo) versus the baseline (youngest) age group. For both sexes there was no evidence that the relationship with age category and having a decreased odds of meeting GHQ-12 cut-off was linear. See Figure 2.

**Figure 2:**
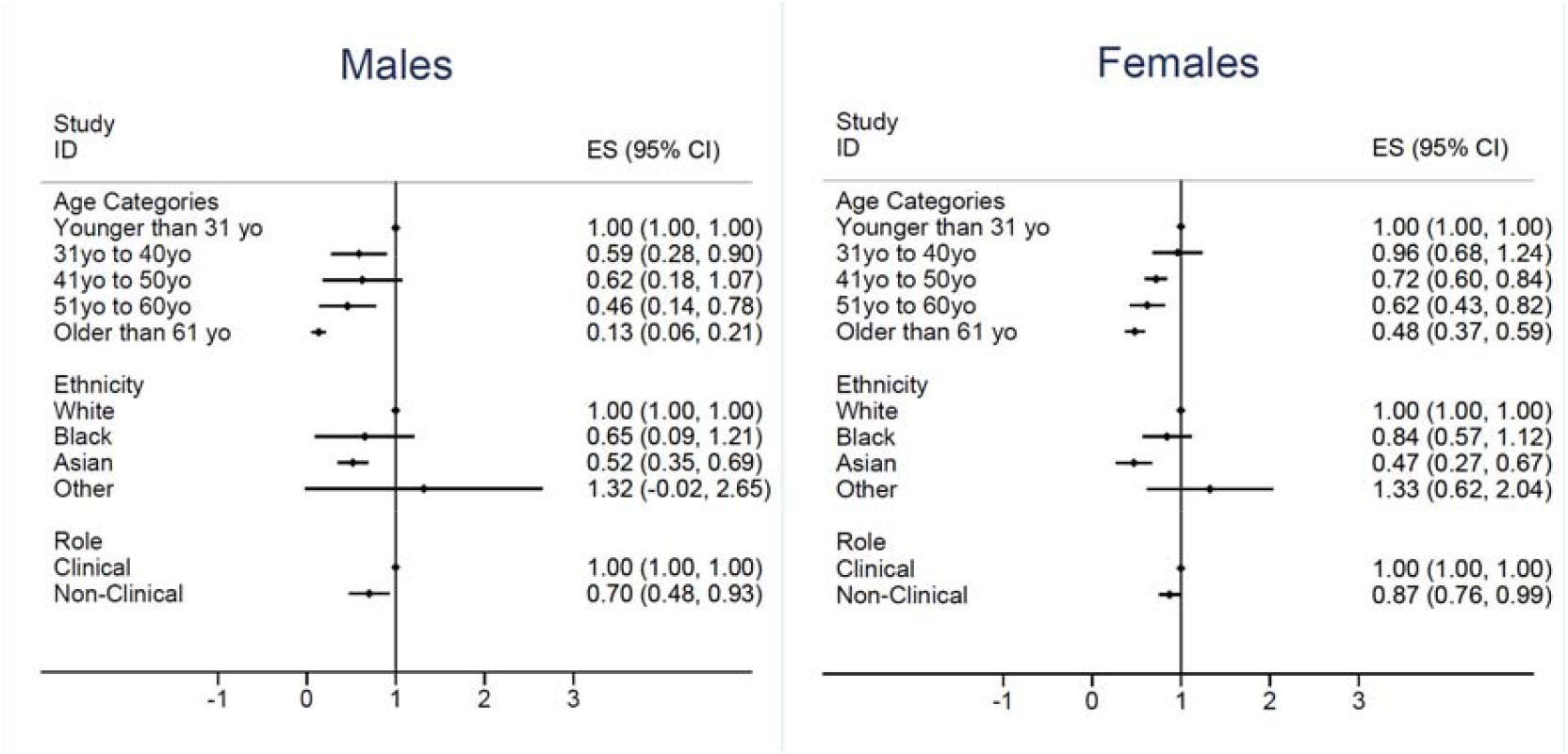
Odds of meeting GHQ cut-off by sex

### Secondary outcomes

The results of similar analyses for the secondary outcomes (anxiety, depression, alcohol misuse, and PTSD) are given in Supplementary figures S1, S2, S3, and S4, respectively.

Similarly to the cross-sectional analyses, older staff of both sexes had statistically significantly decreased odds of meeting GAD-7 cut-off. There were mixed results regarding ethnicity, with males of Other racial or ethnic group having an increase in the odds of meeting cut-off compared to White males, and males and females of Asian ethnicity having reduced odds compared to those of White ethnicity.

Those who were of Asian ethnicity also had a statistically significant lower odds of meeting PHQ-9 cut-off versus those who were White for both sexes. For males, those who were Black had a statistically significantly lower odds of meeting PHQ-9 cut-off versus White participants but not for females. For both sexes, as age increased there was a decrease in the odds of meeting PHQ-9 cut-off.

There were statistically significantly reduced odds of meeting AUDIT cut-off for both sexes of all ethnicities, compared to those of White ethnicity. For both sexes, there was evidence of a decrease in the odd of meeting PCL-6 cut-off as age increased.

## Discussion

### Summary of findings

We followed a national cohort of 22,501 HCW participants, weighted to represent local Trust demographics, and found that prevalence of probable common mental disorders varied over the course of the pandemic, with higher proportions of all staff (clinical and non-clinical) reporting symptoms during periods of increased pressure on the NHS. Prevalence of reported symptoms of anxiety increased over the study period, suggesting that the duration of the pandemic may be taking its toll on overstretched staff.

Younger, female staff, those with inadequate access to PPE and greater exposure to morally injurious events were factors associated with higher prevalence of reported mental health symptoms, however there were no consistent patterns in outcomes terms of ethnicity, job setting, or job role.

### Findings in relation to previous research

The UK Household Longitudinal Study (UKHLS) (a nationally representative probability-based panel study of British adults) found that prevalence of probable CMDs in the general population was statistically significantly higher during the early months of the pandemic (29.5% in April 2020 vs 20.8% in April 2019, 28.% in May 2020, and 26.9% in June 2020), with distress most pronounced for those aged 18-34 and female (Daly & Robinson, 2021). Additional analyses of UKHLS data found no statistically significant increases in GHQ-12 score in keyworkers, comparing data from before/during the pandemic (Pierce, Hope, et al., 2020). Similarly, research comparing professionals from four groups designated as key-workers during the pandemic (i.e. HCW, teachers, public servants, essential services workers) with non-key workers, found no significant differences between the growth trajectories of anxiety and depression symptoms in health and social care workers compared to non-key workers (Paul et al., 2021).

While our results regarding age and sex align with this previous research, we found higher prevalence of distress in our HCW sample (55%, 95%CI 52, 58) than general population studies have done (<30%; Daly & Robinson, 2021). We have previously found that occupational surveys generally report higher prevalence of psychological distress than general population surveys, probably due to a reporting or contextual bias (Goodwin et al., 2013). To counter this, our data were weighed to represent the characteristics of the Trust populations from which our sample was drawn.

The NHS workforce consists of a large proportion of women (76%: NHS Digital, 2021), and our finding that female sex was consistently associated with poorer outcomes at times of increased pressure on the NHS aligns with previous research. Multiple studies have found this both in the UK (Debski et al., 2021; Gilleen et al., 2021; Pappa et al., 2020; Patel et al., 2021; Uphoff et al., 2021; Wanigasooriya et al., 2021) and internationally (De Kock et al., 2021). Qualitative work offers insights into the ingrained pre-pandemic inequalities (e.g. more unpaid caring responsibilities) and asymmetrical power relations, together with gendered organisational structures and norms, which fail to take account of differences (e.g. PPE is frequently made for an average male size) (Regenold & Vindrola-Padros, 2021). However, there are arguments that measures such as the GHQ-12 may not capture the ways in which distress manifests in men (Pierce, Hope, et al., 2020), which may be captured by measures of alcohol use. This bears consideration in light of our finding that men were twice as likely to report alcohol misuse compared with women.

Our results also concur with a small cross-sectional study of UK HCWs from 10 acute and mental health hospitals which also found that younger age and female sex were significantly associated with anxiety and PTSD, and that, in common with our study, inadequate PPE, facing moral dilemmas, and lacking wellbeing support at work were associated with anxiety, depression, and PTSD (Wanigasooriya et al., 2021). Concerns about PPE centred on its quality, and availability and consistency of guidance (Thomas et al., 2020). By following participants over time, we were able to demonstrate that those with good access to PPE were less likely to experience distress in the early months.

Support from colleagues and managers is established as an important factor for workplace wellbeing (Ravalier et al., 2020), as is the deleterious effects of experiencing morally injurious events (Williamson et al., 2021). Being in a higher pay grade (regardless of occupational role) was associated with better outcomes, in line with a wealth of evidence regarding socioeconomic status and mental and physical wellbeing (Marmot et al., 2020). Similarly, as expected those in a relationship had better outcomes (Oskrochi et al., 2018).

However, our hypothesis that ICU staff would be worst affected was supported, with higher prevalence of reported PTSD symptoms across all time periods. Other research also supports the finding of potentially high levels of PTSD in ICU staff (Greenberg et al., 2021). Some research suggests that the majority of HCWs with PTSD symptoms report trauma that pre-dates the pandemic, which may be more likely for those in settings such as ICU because of the nature of their work, however, we have no data on index trauma events (Wild et al., 2021). Counter to our hypothesis around job setting, during the early months of the pandemic, those in A&E were less likely to experience distress than those in all other job settings. This can be explained by the large reduction in A&E use during those months (Leow et al., 2020). We also hypothesised that those redeployed would report worse well-being than those not redeployed, however we found little evidence of this, and at one timepoint (T3), the opposite, with redeployed staff less likely to meet GHQ-12 than non-redeployed staff. Perhaps redeployed staff felt that they were contributing in meaningful ways, which may have counteracted stress associated with moving teams and settings.

Overall, we did not find major or consistent differences in distress between staff in clinical versus non-clinical roles. The exceptions were during summer/autumn of 2020 (T2) and the winter 2020/2021 surge (T3) when doctors (during T2) and nurses (during T3) were more likely than other groups to report distress. Similarly, there were few significant associations between ethnicity and wellbeing outcomes. This may have been due to small numbers, and those most likely to report negative outcomes being less likely to engage with this research. However, these results are surprising, given the discrimination and harassment faced by staff in racial and ethnic minority groups, and the negative effects this has on job satisfaction and sickness absence (Rhead et al., 2021). Alcohol use was the one outcome where there were clear associations with ethnicity: those in Black, Asian, or mixed racial or ethnic groups were less likely to meet cut-off for alcohol misuse than White participants, which is echoed in other research (Niedzwiedz et al., 2021).

### Strengths and limitations

Our study’s major strength is its unprecedented size, comprehensiveness, and likely generalisability: data from over 22,000 staff of all types, with a defined sampling frame of 18 acute and mental health NHS Trusts across England, representing urban and rural areas with differing levels of deprivation. Secondly, the data were weighted according to each participating NHS Trust population’s demographic characteristics, and this means our results can be taken to be representative of those populations. Thirdly, the data were longitudinal, following individuals over time, enabling us to track trends in outcomes. Fourthly, the analyses account for a large number of potential confounders and are multi-level to account for the clustered nature of the data.

There are also some limitations. Firstly, to maximise recruitment and response rate, each data collection wave lasted approximately 10 months, meaning data from individuals were collected at different points in the pandemic, which may have impacted outcomes. We accounted for this by presenting the cross-sectional data partitioned by burden period, but acknowledge that later time periods include follow up data for those recruited at baseline, rather than independent data. Relatedly, the majority of data at T1 came from London-based Trusts, as these were the first recruited, with other Trusts joining towards the end of T1 and during T2. We partially addressed these issues by running longitudinal models that accounted for the date on which measures were completed. Secondly, we do not have pre-pandemic data for our sample, meaning we are unable to assess the role of prior ill-health, nor of the already rising rates of CMDs in younger age groups. Thirdly, while our response rate at baseline was considerably higher than similar studies of HCW wellbeing during the pandemic, at 16% it was lower than we would have liked. Fourthly, to reduce burden on participants (due to known time pressures they would be facing) we split the survey at each time point, with a ‘short’ part that all participants completed, and an optional ‘long’ part that approximately 50% of participants chose to complete in addition. Older, White, and female staff were more likely to complete both the ‘long’ surveys and follow up survey, which may have biased results.

### Implications for future research, policy and practice

Staff in all roles experienced similar levels of distress, which suggests support (whether in terms of practical matters such as staffing and resources, or specific interventions) should not only focus on those in clinical roles and ‘frontline’ staff, but be rolled out across all staff. Flexibility is required, however, as Trusts may need to consider the specific need of certain groups.

Better ways may be needed to reduce the impact of morally challenging situations on staff. Our findings on support from colleagues and managers suggests that ensuring that all NHS staff in supervisory positions are able to competently support their teams, and foster good team working, is likely to lead to better overall staff mental health (Akhanemhe et al., 2021). In turn, leaders and managers need support themselves, and a cascading approach through organisations may be helpful. As the COVID-19 pandemic continues, and in regards to any future pandemics, we expect mental health outcomes to follow a similar pattern, and as such, the need for support may ebb and flow, suggesting careful resource planning in advance is needed. Our evidence suggests that policy early on around PPE in particular could have reduced mental distress, and this should be noted for future planning needs.

## Conclusions

To the best of our knowledge this study provides the best available evidence regarding the mental health and wellbeing of HCWs, with notable strengths including the defined sampling frame, large sample, and longitudinal, weighted data. Our evidence suggests that, contrary to existing research, it is not only clinical staff who are struggling, but all NHS staff. However, there is also some evidence that staff in specific settings such as ICU may be at increased risk. Younger, female, lower paid staff, who feel poorly supported by colleagues and managers, and who experience morally injurious events may be most at risk of negative mental health outcomes, and targeted support for these staff could be particularly helpful.

## Data Availability

Data is available on reasonable request.

## Data sharing statement

Data may be available on request.

## Author contributions

SW, NG, MH, RR, and SAMS are chief investigators of the study, and contributed to manuscript drafts and approved the final draft. DL is a co-investigator of the study, conducted the cross-sectional analyses, and wrote the first draft of the manuscript. RG conducted the longitudinal analyses and contributed to manuscript drafts. EC conducted the weighting of the data and contributed to manuscript drafts. HS and SH contributed to data collection and to manuscript drafts. IM, PM, RM, DM, AMR, SW, and SD are collaborators of the study, and contributed to manuscript drafts.

## Acknowledgments

We wish to acknowledge the National Institute of Health Research (NIHR) Applied Research Collaboration (ARC) National NHS and Social Care Workforce Group, with the following ARCs: East Midlands, East of England, South West Peninsula, South London, West, North West Coast, Yorkshire and Humber, and North East and North Cumbria. They enabled the set-up of the national network of participating hospital sites and aided the research team to recruit effectively during the COVID-19 pandemic.

The NHS CHECK consortium includes the following site leads: Sean Cross, Amy Dewar, Chris Dickens, Frances Farnworth, Adam Gordon, Charles Goss, Jessica Harvey, Nusrat Husain, Peter Jones, Damien Longson, Jesus Perez, Mark Pietroni, Ian Smith, Tayyeb Tahir, Peter Trigwell, Jeremy Turner, Julian Walker, Ashley Wilkie.

The NHS CHECK consortium includes the following co-investigators and collaborators: Peter Aitken, Anthony David, Rosie Duncan, Cerisse Gunasinghe, Stephani Hatch, Daniel Leightley, Isabel McMullen, Martin Parsons, Catherine Polling, Alexandra Pollitt, Danai Serfioti, Chloe Simela, Charlotte Wilson Jones.

## References

Akhanemhe, R., Wallbank, S., & Greenberg, N. (2021). An evaluation of REACTMH mental health training for healthcare supervisors. Occupational Medicine (Oxford, England), 71(3), 127–130. https://doi.org/10.1093/occmed/kqab023

Babor, T. F., Fuente, J. R. D. L., Saunders, J., Grant, M., Babor, T. F., Fuente, J. R. D. L., Saunders, J., Grant, M., & T, H. (2001). The Alcohol Use Disorders Identification Test. World Health Organisation. http://citeseerx.ist.psu.edu/viewdoc/download?doi=10.1.1.505.4146&rep=rep1&type=pdf

Daly, M., & Robinson, E. (2021). Longitudinal changes in psychological distress in the UK from 2019 to September 2020 during the COVID-19 pandemic: Evidence from a large nationally representative study. Psychiatry Research, 300, 113920. https://doi.org/10.1016/j.psychres.2021.113920

Daly, M., Sutin, A., & Robinson, E. (2020). Longitudinal changes in mental health and the COVID-19 pandemic: Evidence from the UK Household Longitudinal Study [Preprint]. PsyArXiv. https://doi.org/10.31234/osf.io/qd5z7

De Kock, J. H., Latham, H. A., Leslie, S. J., Grindle, M., Munoz, S.-A., Ellis, L., Polson, R., & O’Malley, C. M. (2021). A rapid review of the impact of COVID-19 on the mental health of healthcare workers: Implications for supporting psychological well-being. BMC Public Health, 21(1), 104. https://doi.org/10.1186/s12889-020-10070-3

Debski, M., Abdelaziz, H. K., Sanderson, J., Wild, S., Assaf, O., Wiper, A., Nabi, A., Abdelrahman, A., Eichhofer, J., Skailes, G., Gardner, J., Moynes, K., Goode, G., Pathan, T., Patel, B., Kumar, S., Taylor, R., Galasko, G., More, R., … Choudhury, T. (2021). Mental Health Outcomes Among British Healthcare Workers—Lessons From the First Wave of the Covid-19 Pandemic. Journal of Occupational and Environmental Medicine, 63(8), e549–e555. https://doi.org/10.1097/JOM.0000000000002279

Gilleen, J., Santaolalla, A., Valdearenas, L., Salice, C., & Fusté, M. (2021). Impact of the COVID-19 pandemic on the mental health and well-being of UK healthcare workers. BJPsych Open, 7(3). https://doi.org/10.1192/bjo.2021.42

Gnanapragasam, S. N., Hodson, A., Smith, L. E., Greenberg, N., Rubin, G. J., & Wessely, S. (2021). COVID-19 Survey Burden for Healthcare Workers: Literature Review and Audit. Public Health. https://doi.org/10.1016/j.puhe.2021.05.006

Goldberg, D. P., & Hillier, V. F. (1979). A scaled version of the General Health Questionnaire. Psychological Medicine, 9(1), 139–145. https://doi.org/10.1017/S0033291700021644

Goodwin, L., Ben-Zion, I., Fear, N. T., Hotopf, M., Stansfeld, S. A., & Wessely, S. (2013). Are Reports of Psychological Stress Higher in Occupational Studies? A Systematic Review across Occupational and Population Based Studies. PLOS ONE, 8(11), e78693. https://doi.org/10.1371/journal.pone.0078693

Greenberg, N., Weston, D., Hall, C., Caulfield, T., Williamson, V., & Fong, K. (2021). Mental health of staff working in intensive care during Covid-19. Occupational Medicine, 71(2), 62–67. https://doi.org/10.1093/occmed/kqaa220

Kroenke, K., Spitzer, R. L., & Williams, J. B. W. (2001). The PHQ-9. Journal of General Internal Medicine, 16(9), 606–613. https://doi.org/10.1046/j.1525-1497.2001.016009606.x

Lamb, D., Gnanapragasam, S., Greenberg, N., Bhundia, R., Carr, E., Hotopf, M., Razavi, R., Raine, R., Cross, S., Dewar, A., Docherty, M., Dorrington, S., Hatch, S., Wilson-Jones, C., Leightley, D., Madan, I., Marlow, S., McMullen, I., Rafferty, A.-M., … Wessely, S. (2021). Psychosocial impact of the COVID-19 pandemic on 4378 UK healthcare workers and ancillary staff: Initial baseline data from a cohort study collected during the first wave of the pandemic. Occupational and Environmental Medicine, 78(11), 801–808. https://doi.org/10.1136/oemed-2020-107276

Lamb, D., Greenberg, N., Hotopf, M., Raine, R., Razavi, R., Bhundia, R., Scott, H., Carr, E., Gafoor, R., Bakolis, I., Hegarty, S., Souliou, E., Rafferty, A. M., Rhead, R., Weston, D., Gnangapragasam, S., Marlow, S., Wessely, S., & Stevelink, S. (2021). NHS CHECK: Protocol for a cohort study investigating the psychosocial impact of the COVID-19 pandemic on healthcare workers. BMJ Open, 11(6), e051687. https://doi.org/10.1136/bmjopen-2021-051687

Lamb, D., Greenberg, N., Stevelink, S. A. M., & Wessely, S. (2020). Mixed signals about the mental health of the NHS workforce. The Lancet Psychiatry, 7(12), 1009–1011. https://doi.org/10.1016/S2215-0366(20)30379-5

Lang, A. J., & Stein, M. B. (2005). An abbreviated PTSD checklist for use as a screening instrument in primary care. Behaviour Research and Therapy, 43(5), 585–594. https://doi.org/10.1016/j.brat.2004.04.005

Leow, S. H., Dean, W., MacDonald-Nethercott, M., MacDonald-Nethercott, E., & Boyle, A. A. (2020). The Attend Study: A Retrospective Observational Study of Emergency Department Attendances During the Early Stages of the COVID-19 Pandemic. Cureus, 12(7), e9328. https://doi.org/10.7759/cureus.9328

Marmot, M., Allen, J., Boyce, T., Goldblatt, P., & Morrison, J. (2020). Marmot Review 10 Years On. Institute of Health Equity. http://www.instituteofhealthequity.org/resources-reports/marmot-review-10-years-on

Marvaldi, M., Mallet, J., Dubertret, C., Moro, M. R., & Guessoum, S. B. (2021). Anxiety, depression, trauma-related, and sleep disorders among healthcare workers during the COVID-19 pandemic: A systematic review and meta-analysis. Neuroscience & Biobehavioral Reviews, 126, 252–264. https://doi.org/10.1016/j.neubiorev.2021.03.024

Nash, W. P., Marino Carper, T. L., Mills, M. A., Au, T., Goldsmith, A., & Litz, B. T. (2013). Psychometric Evaluation of the Moral Injury Events Scale. Military Medicine, 178(6), 646–652. https://doi.org/10.7205/MILMED-D-13-00017

NHS. (2020). Agenda for change—Pay rates. Health Careers. https://www.healthcareers.nhs.uk/working-health/working-nhs/nhs-pay-and-benefits/agenda-change-pay-rates

NHS Digital. (2021). NHS Workforce Statistics—September 2021. https://digital.nhs.uk/data-and-information/publications/statistical/nhs-workforce-statistics/september-2021

NHS England. (2021). NHS staff survey 2021. https://www.nhsstaffsurveys.com/static/afb76a44d16ee5bbc764b6382efa1dc8/ST20-national-briefing-doc.pdf

Niedzwiedz, C. L., Green, M. J., Benzeval, M., Campbell, D., Craig, P., Demou, E., Leyland, A., Pearce, A., Thomson, R., Whitley, E., & Katikireddi, S. V. (2021). Mental health and health behaviours before and during the initial phase of the COVID-19 lockdown: Longitudinal analyses of the UK Household Longitudinal Study. J Epidemiol Community Health, 75(3), 224–231. https://doi.org/10.1136/jech-2020-215060

Office for National Statistics. (2022). Deaths involving COVID-19 by month of registration, UK. Office for National Statistics. https://www.ons.gov.uk/peoplepopulationandcommunity/birthsdeathsandmarriages/deaths/datasets/deathsinvolvingcovid19bymonthofregistrationuk/current

Oskrochi, G., Bani-Mustafa, A., & Oskrochi, Y. (2018). Factors affecting psychological well-being: Evidence from two nationally representative surveys. PLOS ONE, 13(6), e0198638. https://doi.org/10.1371/journal.pone.0198638

Pappa, S., Ntella, V., Giannakas, T., Giannakoulis, V. G., Papoutsi, E., & Katsaounou, P. (2020). Prevalence of depression, anxiety, and insomnia among healthcare workers during the COVID-19 pandemic: A systematic review and meta-analysis. Brain, Behavior, and Immunity, 88, 901–907. https://doi.org/10.1016/j.bbi.2020.05.026

Patel, K., Robertson, E., Kwong, A. S. F., Griffith, G. J., Willan, K., Green, M. J., Gessa, G. D., Huggins, C. F., McElroy, E., Thompson, E. J., Maddock, J., Niedzwiedz, C. L., Henderson, M., Richards, M., Steptoe, A., Ploubidis, G. B., Moltrecht, B., Booth, C., Fitzsimons, E., … Katikireddi, S. V. (2021). Psychological Distress Before and During the COVID-19 Pandemic: Sociodemographic Inequalities in 11 UK Longitudinal Studies (p. 2021.10.22.21265368). https://doi.org/10.1101/2021.10.22.21265368

Paul, E., Mak, H. W., Fancourt, D., & Bu, F. (2021). Comparing mental health trajectories of four different types of key workers with non-key workers: A 12-month follow-up observational study of 21,874 adults in England during the COVID-19 pandemic (p. 2021.04.20.21255817). https://doi.org/10.1101/2021.04.20.21255817

Pierce, M., Hope, H., Ford, T., Hatch, S., Hotopf, M., John, A., Kontopantelis, E., Webb, R., Wessely, S., McManus, S., & Abel, K. M. (2020). Mental health before and during the COVID-19 pandemic: A longitudinal probability sample survey of the UK population. The Lancet Psychiatry, 0(0). https://doi.org/10.1016/S2215-0366(20)30308-4

Pierce, M., McManus, S., Jessop, C., John, A., Hotopf, M., Ford, T., Hatch, S., Wessely, S., & Abel, K. M. (2020). Says who? The significance of sampling in mental health surveys during COVID-19. The Lancet Psychiatry, 7(7), 567–568. https://doi.org/10.1016/S2215-0366(20)30237-6

Preti, E., Di Mattei, V., Perego, G., Ferrari, F., Mazzetti, M., Taranto, P., Di Pierro, R., Madeddu, F., & Calati, R. (2020). The Psychological Impact of Epidemic and Pandemic Outbreaks on Healthcare Workers: Rapid Review of the Evidence. Current Psychiatry Reports, 22(8), 43. https://doi.org/10.1007/s11920-020-01166-z

Ravalier, J. M., McVicar, A., & Boichat, C. (2020). Work Stress in NHS Employees: A Mixed-Methods Study. International Journal of Environmental Research and Public Health, 17(18), 6464. https://doi.org/10.3390/ijerph17186464

Regenold, N., & Vindrola-Padros, C. (2021). Gender Matters: A Gender Analysis of Healthcare Workers’ Experiences during the First COVID-19 Pandemic Peak in England. Social Sciences, 10(2), 43. https://doi.org/10.3390/socsci10020043

Rhead, R. D., Chui, Z., Bakolis, I., Gazard, B., Harwood, H., MacCrimmon, S., Woodhead, C., & Hatch, S. L. (2021). Impact of workplace discrimination and harassment among National Health Service staff working in London trusts: Results from the TIDES study. BJPsych Open, 7(1). https://doi.org/10.1192/bjo.2020.137

Scavone, F. (2021). Pay scales for junior doctors in England. The British Medical Association Is the Trade Union and Professional Body for Doctors in the UK. https://www.bma.org.uk/pay-and-contracts/pay/junior-doctors-pay-scales/pay-scales-for-junior-doctors-in-england

Smith, R. (2020). Earnings and working hours 2020. Office for Natonal Statistics. https://www.ons.gov.uk/employmentandlabourmarket/peopleinwork/earningsandworkinghours/bulletins/annualsurveyofhoursandearnings/2020

Spitzer, R. L., Kroenke, K., Williams, J. B. W., & Löwe, B. (2006). A Brief Measure for Assessing Generalized Anxiety Disorder: The GAD-7. Archives of Internal Medicine, 166(10), 1092–1097. https://doi.org/10.1001/archinte.166.10.1092

Thomas, J. P., Srinivasan, A., Wickramarachchi, C. S., Dhesi, P. K., Hung, Y. M., & Kamath, A. V. (2020). Evaluating the national PPE guidance for NHS healthcare workers during the COVID-19 pandemic. Clinical Medicine, 20(3), 242–247. https://doi.org/10.7861/clinmed.2020-0143

Uphoff, E. P., Lombardo, C., Johnston, G., Weeks, L., Rodgers, M., Dawson, S., Seymour, C., Kousoulis, A. A., & Churchill, R. (2021). Mental health among healthcare workers and other vulnerable groups during the COVID-19 pandemic and other coronavirus outbreaks: A rapid systematic review. PLOS ONE, 16(8), e0254821. https://doi.org/10.1371/journal.pone.0254821

Wanigasooriya, K., Palimar, P., Naumann, D. N., Ismail, K., Fellows, J. L., Logan, P., Thompson, C. V., Bermingham, H., Beggs, A. D., & Ismail, T. (2021). Mental health symptoms in a cohort of hospital healthcare workers following the first peak of the COVID-19 pandemic in the UK. BJPsych Open, 7(1). https://doi.org/10.1192/bjo.2020.150

Wild, J., McKinnon, A., Wilkins, A., & Browne, H. (2021). Post-traumatic stress disorder and major depression among frontline healthcare staff working during the COVID-19 pandemic. British Journal of Clinical Psychology. https://doi.org/10.1111/bjc.12340

Williamson, V., Murphy, D., Phelps, A., Forbes, D., & Greenberg, N. (2021). Moral injury: The effect on mental health and implications for treatment. The Lancet Psychiatry, 8(6), 453–455. https://doi.org/10.1016/S2215-0366(21)00113-9

Williamson, V., Stevelink, S. A. M., & Greenberg, N. (2018). Occupational moral injury and mental health: Systematic review and meta-analysis. The British Journal of Psychiatry, 212(6), 339–346. https://doi.org/10.1192/bjp.2018.55

